# Covid-19 Risk Among Airline Passengers: Should the Middle Seat Stay Empty?

**DOI:** 10.1101/2020.07.02.20143826

**Authors:** Arnold Barnett, Keith Fleming

## Abstract

We use recent data and research results and a probabilistic model to estimate the chance that an air traveler in coach will contract Covid-19 on a US domestic jet flight two hours long, both when all coach seats are full and when all but middle seats are full. The point estimates we reach based on data from late September 2020 are about 1 in 3,900 for full flights and 1 in 6,400 when middle seats are kept empty. These estimates are subject to substantial uncertainty, with factor-of-three or greater margins of error. However, because uncertainties in key parameters affect both risk estimates the same way, they leave the relative risk ratio for “fill all seats” compared to “middle seat open” close to 1.64 (i.e., close to (1/3,900)/(1/6,400). We compare the infection risks over a two-hour flight to those of two hours on the ground, and find that the flight presents greater hazard. We also approximate the mortality risks caused by Covid-19 infections contracted on airplanes, taking into account that infected passengers can in turn infect others not on the plane. The point estimates for death risk are low—averaging about one death per 800,000 passengers--but they are somewhat higher than those associated with plane crashes and aviation terrorism.

## Introduction

The Covid-19 crisis has led to something unprecedented: a public disagreement among US airlines on a question of safety. As of October 2020, Alaska, Delta, jetBlue, and Southwest Airlines are keeping middle seats open on their flights to limit infection risk, while Allegiant, American, Spirit, and United Airlines are selling all seats when demand warrants. United Airlines has vigorously defended its policy, describing “middle seats open” as a “PR strategy and not a safety strategy.” In contrast, Delta Airlines CEO Edward Bastian pointedly stated that the carrier doesn’t fill middle seats because it “puts safety before profits.”

Outside the industry, prominent experts have expressed dismay at the “fill all seats” policy. When American Airlines announced that it would sell as many seats as it could, Dr. Anthony Fauci, the top infectious diseases official at the US National Institutes for Health, told a Senate hearing that “obviously, that’s something that is of concern.” Dr. Robert Redfield, the director of the US Centers for Disease Control and Prevention (CDC), agreed, declaring that “I can tell you that when they announced that the other day, obviously there was substantial disappointment with American Airlines.”

Approximately 80 million passengers flew on US domestic flights between June and September 2020. One might therefore imagine that we would have direct information by now about how often passengers with contagious Covid-19 board US flights, and about how much transmission has occurred aboard aircraft as a function of mask usage and the level of crowding. After all, even under the improbably-low assumption that 1 in 2000 of the boarding passengers were Covid-19 positive, that would work out to 40,000 infected people. The evidence is strong, however, that few of those individuals have been identified and, even for those who have been, contract tracing for passengers seated close to them has been sparse at best. Under the circumstances, the fact that no “confirmed” cases of on-board infection are at hand cannot be taken too seriously: an official from the US Centers for Disease Control (CDC) pointed out that “an absence of cases identified or reported is not evidence that there were no cases” [1]. An extensive review of available studies reached the same conclusion, namely, that “the absence of large numbers of published in-flight transmissions of SARS-CoV-2 is not definitive evidence of safety” [2].

There are trustworthy reports about coronavirus transmission aboard some recent international flights, but these flights differ appreciably from those in US domestic service, especially in terms of duration and the role of masks. Against that backdrop, it becomes important to develop a probabilistic model to approximate the Covid-19 risks to travelers on US domestic jet flights. This paper is an attempt to do so, with special reference to the divergence between “middle seats empty” and “fill all seats” policies. The estimation requires a variety of assumptions, on such topics as the likelihood that a boarding passenger carries contagious Covid-19, the effectiveness of the masks that passengers typically use, and the risk of successful transmission absent masks. The endeavor is imperfect, but it seems preferable to a bitter clash of conjectures.

Using data from late September 2020 and earlier research findings, we make the first-order approximation that, on full flights two hours long on popular US jets (the Boeing 737 and the Airbus 320), coach passengers sustain approximately a 1 in 3900 risk of contracting Covid-19 from a nearby passenger. Under “middle seat empty,” the risk is approximately 1 in 6400, a factor of 1.64 lower. Both these estimates are subject to sizable uncertainty, though the factor of 1.64 is considerably less so. Given these estimates and some others, cases of Covid-19 contracted aboard domestic flights could directly or indirectly cause one approximately one death per 710,000 passengers on flights by airlines that would sell all seats if they could (and that actually filled 85% of seats). Under “middle seat empty,” the corresponding figure is about one death per 920,000 passengers.

We start our work in Section II, where we introduce the general model for assessing Covid-19 risk for uninfected passengers on two-hour flights. (We focus on two hours because that is the approximate duration of an average US jet flight.). In Section III, we describe the probability distributions we assign to seven key parameters. Then in Section IV, we present the results of a Monte Carlo simulation that generates a probability distribution for Covid-19 risk for a randomly-chosen uninfected passenger. We discuss the results in Section V and offer some concluding remarks in Section VI.

### II. A General Model

The model of on-board Transmission Risk from a contagious person follows the basic equation:

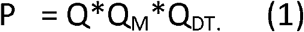

Where P = unconditional probability that an uninfected traveler on a US domestic jet flight two hours long comes down with Covid-19 during the flight

Q = the expected number of contagious travelers on such a US domestic flight

Q_M_ = an estimate of the probability that passenger masks (universally required on such flights), *fail* to prevent the transmission of infection, despite the effectiveness of masks in protecting the wearer from others and protecting others from the wearer

Q_DT_ = a factor that reflects how, absent masks and conditioned on contagious passenger(s), transmission risk depends on both the duration of the flight and the seating locations of both the contagious passenger(s) and other travelers

### The Estimation of Q

Q is given by:

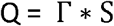

S = number of passengers on the flight

Γ = probability that a given passenger boards with a contagious Covid-19 infection

We model Γ as a product of several factors:

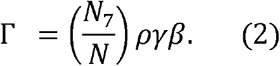

Where: *N*_7_ = Number of confirmed cases of Covid – 19 in the region of interest over the past seven days

*N* = Population of region of interest

*ρ =* a multiplier to reflect the extent to which confirmed Covid – 19 cases underestimate the actual number of cases

*γ* = a factor to reflect the proportion of Covid-19 individuals who are asymptomatic, as well as the research finding that asymptomatic carriers of the diseases are less likely to be contagious than pre-symptomatic or symptomatic carriers

*β* = a “healthy passenger” factor to reflect the likelihood that the per capita rate of Covid - 19 among travelers boarding airplanes is lower than that in the population as a whole.

Equation (2) is based on the evidence that (i) the number of actual cases of Covid-19 in the US is a large multiple of the number of confirmed cases, (ii) asymptomatic carriers of the disease are considerably less contagious than pre-symptomatic and symptomatic ones, (iii) air travelers are considerably less likely to be contagious than the citizenry as a whole.

The quantity N_7_ enters (2) because seven days is the approximate length of the contagiousness period for someone experiencing Covid-19. (The average such period is a bit below seven days in asymptomatic cases and higher than seven in symptomatic ones; see [3,4].) Dividing N_7_ by N, the population of the region of interest, yields the region’s per capita rate of new *confirmed* cases over the last week.

The factor *ρ* arises because of recognition that only a fraction of Covid-19 infections get confirmed in the US. In a paper that appeared in August 2020, CDC researchers offered evidence that infections over March-May 2020 were underestimated by a factor of ten [5]. More recently, underestimation has apparently diminished, but by no means has it disappeared.

People with Covid-19 who board airplanes are asymptomatic, pre-symptomatic, or symptomatic. Researchers have estimated the proportion of Covid-19 carriers who are asymptomatic, and offered evidence that carriers who are asymptomatic are less contagious than others. The factor *γ* “discounts” the risk estimate to take account of the lesser risk posed by infected but asymptomatic passengers.

It is known that air travelers are not randomly spread over the population: they are more likely to be affluent, to be able to avoid catching Covid-19 because of their jobs, and to live in communities that have relatively low confirmed rates of Covid-19 per capita (excluding nursing homes, almost none of whose residents are flying during the pandemic.). For that reason the “healthy traveler” multiplier *β* is important to the risk calculation, and is probably well below one.

### The Quantities Q_M_ and Q_DT_

The analysis here depends substantially on two recent papers:

- *A 2020 meta-analysis in *The Lancet* [6], which considered 216 studies, and which estimated the effectiveness of masks against viral infections and the dependence of infection risk on the distance between the contagious person and the uninfected one.
- A 2018 study in *Proceedings of the National Academy of Sciences* (PNAS) [7], which explored how the probability of viral infection on an aircraft depends on the proximity and duration of exposure to a contagious person

The *Lancet* paper and other articles generate a probability distribution for how successfully the cloth masks and surgical masks that US air travelers generally use prevent the transmission of viral emissions from a Covid-19 contagious person. They thus offer a distribution for the complementary probability Q_M_ that the masks fail to prevent transmission.

The estimation of Q_DT_ must account for the fact that the chance that a passenger with contagious Covid-19 will infect another passenger depends on both the duration of the flight and the locations of the two travelers within the aircraft. But estimating the transmission probability (absent masks) is very difficult. It depends on the contagious passenger’s emissions of the virus via a mixture of breathing, speaking, and coughing or sneezing (a mixture that varies from person to person), as well as the movement of droplets and aerosols given the geometry of the airplane and its powerful HEPA air-purification systems. None of the processes is fully understood for Covid-19.

There has, however, been a direct attempt to estimate viral-transmission probabilities aboard an airplane, namely, the PNAS study cited above. The authors approximated the transmission probability as 1.8% for each minute the uninfected traveler is within one meter of the contagious one and no masks are in use. They assumed that the viral output from the contagious passenger was “omnidirectional,” and they stated that they did not consider the possibility that seatbacks served as barriers to transmission. The authors focused on US transcontinental flights about four hours long, and therefore estimated the chance that a passenger seated within one meter of a contagious traveler would become infected as approximately 1 - .982^240^ = .987. In all, the authors projected that one contagious passenger could generate roughly 12 infections over the course of a flight with all seats full.

Here we do not use on the parameter estimate of .018 per minute (which the authors say they had inflated by a factor of four to be conservative). But we will follow the PNAS paper in making the approximation that, on a two-hour flight without mask usage, the chance H_D_ that a given uninfected passenger contracts Covid-19 from a particular contagious passenger takes the general form:

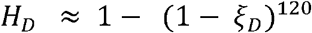

Where *ξ*_*D*_ = transmission probability per minute given the relative positions of the contagious and uninfected passengers.

While the PNAS paper breaks distances from the contagious passenger with the dichotomy “below one meter/above one meter,” we follow the *Lancet paper in treating* contagion risk as a continuous function of distance. In particular, we adopt the conclusion from their meta-analysis that, absent barriers between the contagious and uninfected passenger, risk decays exponentially with distance. However, we allow for the likelihood that seatbacks constitute transmission barriers that, while not as effective as plexiglass, do block some emissions from a given row to adjacent rows.

Given these considerations, we posit that *ξ*_*D*_ takes the general form

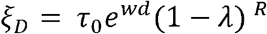

Where d = distance between the contagious and the uninfected passenger

*τ*_0_ = the transmission probability per minute to someone zero distance from the contagious passenger (or the chance per minute that that passenger would “self-infect” if that were possible)

*ω* = the (negative) parameter that specifies the rate of exponential decay of risk with distance

*λ* = the effectiveness of the seatback as a barrier, relative to plexiglass (*λ* < 1)

R = number of seatbacks separating the contagious and the uninfected passenger (e.g., between seats 16B and 14A, R=2; between 14C and 14A, R = 0)

(As noted: *ξ*_*D*_ is conditioned on both a contagious passenger on board and the failure of masks.)

In measuring d, we use not the Euclidean distance between the contagious passenger and others but instead the “grid distance” (i.e., d((x_1_, y_1_) to (x_2,_ y_2_)) = |x_2_ – x_1_| + |y_2_ – y_1_|). This choice reflects the assumption that emissions from a contagious traveler in 16A that reach the breathing space of a passenger in 15B could well travel there via the breathing space for the passenger in 15A.

A contagious passenger can simultaneously infect many others. *H*_*DT*_, that person’s expected number of transmissions (without masks) follows the rule:

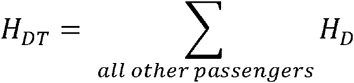

We will argue that, as a practical matter, the exponential decay of risk with distance and the growing number of seatback-barriers mean that the risk to passengers more than two rows from the contagious traveler are second-order effects. We will also argue that transmission risk for a randomly-selected uninfected traveler is generally *H*_*DT*_ divided by S_NI_, where S_NI_ is the number of uninfected passengers aboard the flight

Putting it all together, the Covid-19 risk model takes the general form:

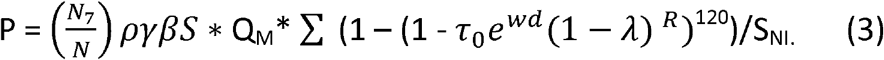

### III. Probability Distributions for Model Parameters

In all, there are seven distinct parameters in (3) that are not known exactly, namely, *ρ, γ, and β,Q*_*M*_, *τ*_0_,*ω and λ*. We assign probability distributions to each of them, recognizing that doing so is imperfect given uncertainties about the Covid-19 epidemic in the United States.

#### Distributions Related to Q

The unknown parameters in the expression for Q in (2) are *ρ, γ, and β*, while N and N_7_ are known. Here we concentrate on the US as a whole and the week from September 17 to 23 in 2020. The official statistic is that 253,000 US cases of Covid-19 were confirmed over 9/17/20 through 9/23/20 (i.e., N_7_ was 253,000.) [8], while the US population (N) was about 330 million.

The quantity *ρ* concerns the large disparity between confirmed cases of Covid-19 and the actual number of cases. Researchers at the Centers for Disease Control (CDC) estimated that there were approximately nine unconfirmed cases in the US over March-May 2020 for every case that was confirmed [5] But there are indications that the greater availability of Covid-19 testing has considerably reduced the undercount of actual cases. The question is how much.

We can approximate *ρ* through a form of “reverse engineering.” It was estimated in June 2020 that 0.7% of Covid-19 infections in the US ended in death [9], which works out to about one death per 141 cases (the case fatality rate). Thus, multiplying reported deaths in a given period by 141 would yield an approximate number for new cases some weeks earlier, taking account of the time lag between infection and deaths. However, recent improvements in treatment for Covid-19 are believed to have lowered the case fatality rate in recent months by perhaps 20-33%, to about 0.5% [10,11],. Thus, multiplying by 200 (i.e., 1/.005) could be more accurate. Once estimated numbers of cases are at hand, they can be compared to confirmed cases to yield estimates of *ρ*.

Table 1 presents estimates of *ρ* based on this approach for the three weeks in September 2020, assuming intervals of two to five weeks between confirmed infection and death. These ratios are quite stable over that period, averaging about 2.2 based on 141 infections per death and 3.1 based on 200. But given that this estimation method is itself approximate, it seems prudent to assign *ρ* a uniform distribution on the range from 1.5 to 4. We do so here.

**Table 1:**
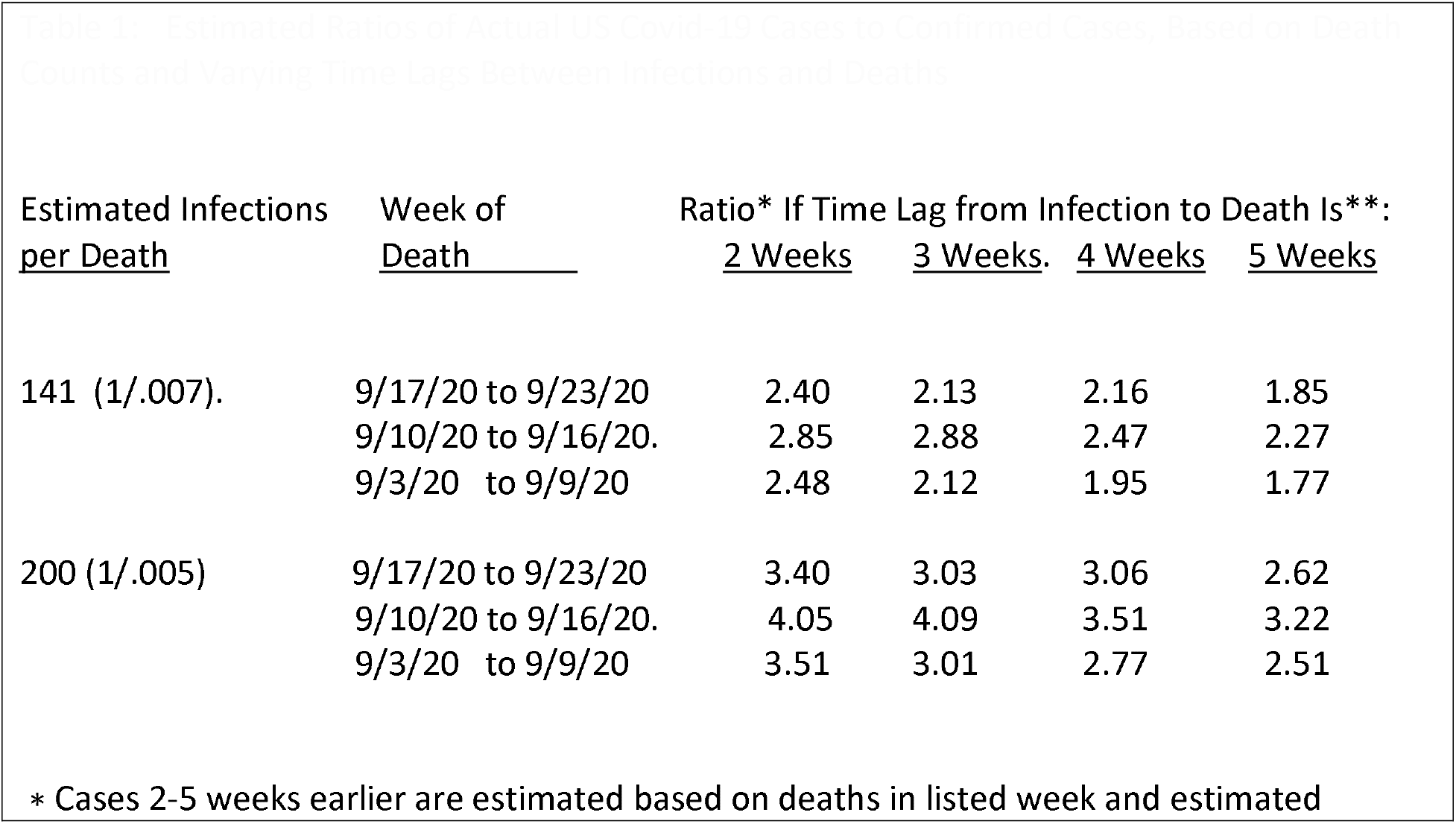
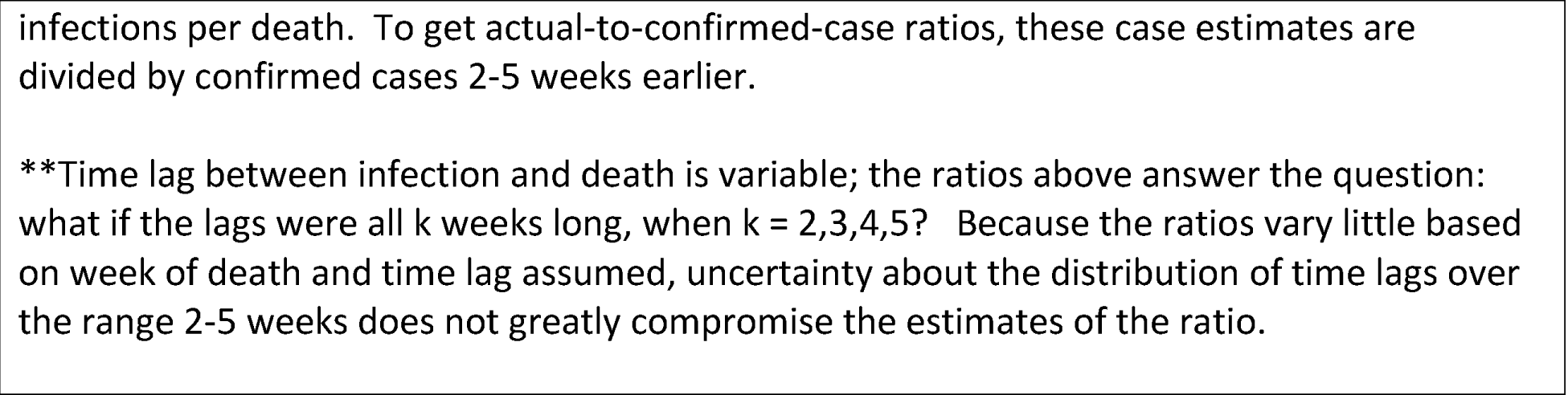
Estimated Ratios of Actual US Covid-19 Cases to Confirmed Cases, Based on Death Counts and Varying Time Lags Between Infections and Deaths

| <u>Estimated Infections</u><br><u>per Death</u> | <u>Week of</u><br><u>Death</u> | Ratio* If Time Lag from Infection to Death Is**: |  |  |  |
| --- | --- | --- | --- | --- | --- |
|  |  | <u>2 Weeks</u> | <u>3 Weeks.</u> | <u>4 Weeks</u> | <u>5 Weeks</u> |
| 141 (1/.007). | 9/17/20 to 9/23/20 | 2.40 | 2.13 | 2.16 | 1.85 |
|  | 9/10/20 to 9/16/20. | 2.85 | 2.88 | 2.47 | 2.27 |
|  | 9/3/20 to 9/9/20 | 2.48 | 2.12 | 1.95 | 1.77 |
| 200 (1/.005) | 9/17/20 to 9/23/20 | 3.40 | 3.03 | 3.06 | 2.62 |
|  | 9/10/20 to 9/16/20. | 4.05 | 4.09 | 3.51 | 3.22 |
|  | 9/3/20 to 9/9/20 | 3.51 | 3.01 | 2.77 | 2.51 |

\* Cases 2-5 weeks earlier are estimated based on deaths in listed week and estimated
infections per death. To get actual-to-confirmed-case ratios, these case estimates are divided by confirmed cases 2-5 weeks earlier.
**\*\*Time lag between infection and death is variable; the ratios above answer the question: what if the lags were all k weeks long, when k = 2,3,4,5? Because the ratios vary little based on week of death and time lag assumed, uncertainty about the distribution of time lags over the range 2-5 weeks does not greatly compromise the estimates of the ratio.**

The factor *γ* is based on both the fraction of Covid-19 cases that are asymptomatic and the reduced contagiousness of asymptomatic carriers. If Z_1_ the asymptotic proportion of cases (Z_1_) and Z_2_ is asymptotic infectiousness (Z_2_) then the probability *γ* that a randomly-chosen person infected with Covid-19 is contagious is given by *γ* = (1-z_1_)+ z_1_z_2_. (The first term on the right is the chance the person is symptomatic (and thus assumed to be contagious), while the second is the chance the person is both asymptomatic and contagious.) Research suggests that 40% to 45% of individuals with Covid-19 are asymptomatic [12], and that such individuals are 25% to 66% as likely to infect others as symptomatic carriers of the disease [13}. Given these findings, we model Z_1_ and Z_2_ as independent normal random variables, with *z*_1_ ≈ *N*(.4,.05) *and z*_2_ ≈ *N*(.45,.1)

Of the three multipliers that appear in Q, the most difficult to estimate is *β*, which approximates the extent to which travelers boarding US jet flights are less likely to be infected with Covid-19 than randomly-chosen US residents. Suppose that boarding passengers are randomly spread among the half of Americans less likely to carry Covid-19, and that X, the per capita rate of Covid-19 within this half of Americans, is ¼ as high as that in the riskier half. Then the average disease rate among Americans would be 2.5X (i.e., (X + 4X)/2), and the risk multiplier *β* for passengers would be X/2.5X = 0.4. If the disease rate in the riskier half of Americans is 2X, the corresponding multiplier would be 0.667; if that rate were 6X, the multiplier would be 0.283. Here we assign *β* a triangular distribution:

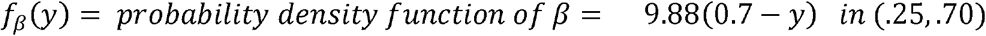

This density function has a mean of .40 and, within the range from .25 to 70, it assigns higher probabilities to greater levels of disparity. Direct empirical evidence about the value of *β* is lacking now.

#### Distribution of QM

Some passengers on US domestic flights use cloth masks, while others wear surgical masks. The meta-analysis in the Lancet paper estimates that wearing of cloth or surgical masks reduces by an estimated 67% the chance that a viral infection is successfully transmitted. The confidence interval for the benefit extends from 39% to 83% A study from China [14] about Covid-19 yields the estimate that, when everyone wears such masks, overall infection risk drops 79%. Another paper about viral infections [15} estimates both a 50% reduction of the risk of outward transmission by a cloth-mask wearer and 50% drop in the risk of inward transmission. Treating these two benefits as independent implies that universal cloth-mask wearing would cut transmissions by 75%. (i.e. to 50%*50% of the level with no masks). The corresponding reduction for surgical masks was estimated as 94%. Giving by far the greatest weight to the meta-analysis, we approximate the risk multiplier *Q*_*M*_ as approximately normal with a mean of 0.3 (i.e., a 70% reduction) and a standard deviation of 0.1.

#### Distributions Related to QDT

The three parameters in Q_DT_ are *τ*_0_,, *ω, and λ*, which enter expressions of the general form (1 − *τ*_0_*e*^*wd*^(1 − *λ*)^*R*^)^120^ Assigning a distribution to is easier than doing so for *τ*_0_ *and λ*

As noted, the Lancet meta-analysis estimated that viral transmission risk declines exponentially with greater distance from the contagious person. The point estimate was that risk declined by a factor of 2.02 for each additional meter of separation, with a 95% confidence interval extending from a factor of 1.08 to one of 3.76. This interval—from roughly half the point estimate to double it—is strongly suggestive of a lognormal distribution.

Because ln(1/2.02) = -.703, ln(1/1.08) = -.077, and ln(1/3.76) = −1.324, we assign the logarithm of the decay parameter *ω* a mean of -.703 and a standard deviation of .318. (The latter statistic is (1/1.96) times the average of 1.324 - .703 = .621 and .703-.077= .626.)

In each coach row in a typical Boeing 737 or an Airbus 320, the individual seats are approximately 18 inches wide, while the aisle width is about 30 inches. The seatbacks in consecutive rows are separated by about 31 inches. With these dimensions, the grid distances in inches from people within two rows of a contagious person in seat 16A are:

| Seat |  |  |  |  |  |  |
| --- | --- | --- | --- | --- | --- | --- |
| Row | A | B | C | D | E | F |
| 14 | 62 | 80 | 98 | 128 | 146 | 164 |
| 15 | 31 | 49 | 67 | 97 | 115 | 133 |
| 16 | 0 | 18 | 36 | 66 | 84 | 102 |
| 17 | 31 | 49 | 67 | 97 | 115 | 133 |
| 18 | 62 | 80 | 98 | 128 | 146 | 164 |

One can create similar charts when the contagious passenger is in a B or C seat and, by symmetry, in the D, E, or F seat.

Passengers three or more rows from the contagious person are separated from him by a minimum of 91 inches and three seatbacks. For that reason, we treat their infection risks as second-order effects. We further assume that short interactions among passengers, during boarding, deplaning, or en route to the lavatory are also second-order effects in the risk estimates. We make the same assumption for the short periods when passengers take off their masks to consume drinks or foods. In this connection, we lean upon the CDC guidance that a person exposed to someone with confirmed Covid-19 should go into quarantine if the interaction was at least 15 minutes long. The interactions just mentioned are typically far shorter than 15 minutes. If, however, one believes that these hazards are not negligible, they would add to the risks that we are estimating.

As barriers to the spread of Covid-19, seatbacks are inferior to floor-to-ceiling shields of plexiglass. Seatbacks cannot prevent a viral emission from passing above or below them. Still, the seatback directly ahead of a disease sufferer can block some forward transmission, and that person’s own seatback can somewhat protect the passengers one row behind. Lacking literature about the health benefits of seatbacks, we assign *λ*, the ratio of setback effectiveness to that of plexiglass, a uniform distribution between 0.3 and 0.8. Then 1 - *λ* is the failure rate of seatbacks relative to plexiglass, which we treat as a perfect barrier if floor-to-ceiling.

The estimation of *τ*_0_ -the transmission risk per minute of exposure absent masks and zero distance from the contagious passenger--is especially challenging. We do so having considered the following research papers, which generally refer to flights outside the United States. Corresponding data for US domestic flights, especially with reference to Covid-19, are essentially unavailable.

1. Hertzberg and Weiss wrote an influential paper that discussed the spread of SARS in 2003 on a flight from Hong Kong to Beijing [16]. Nine of the 23 passengers within two rows of the contagious passenger came down with SARS, as did nine of those more than two rows away.
2. Khanh et. al. analyzed the spread of Covid-19 aboard Vietnam Airlines Flight 54 from London to Ho Chi Minh City [17], which was about ten hours and ten minutes long. They report that a single contagious passenger in Business class infected 12 of the other 21 Business class passengers, including 11 of the 12 within two meters of the contagious traveler and 1 of 8 more than two meters away. There is no indication that any of the passengers wore masks.
3. Hoehl et. al. studied infection patterns on a four hour and 40 minute flight from Tel Aviv to Frankfurt, in which seven passengers seated together boarded with Covid-19 infections (i.e. had *primary* infections) and likely transmitted the disease to two of the twelve passengers seated nearby (i.e. who got secondary infections) [18]. Again, apparently no masks were worn.
4. Speakes et. al. report that, on a five-hour flight from Sydney to Perth on which masks were not used, eight passengers with primary Covid-19 infections were scattered throughout the coach section [19]. They collectively generated eight secondary Covid-19 infections among 20 passengers within two rows of them.
5. Moser et. al. reported on a flight in Alaska in which 38 of the 54 passengers seemingly got secondary influenza infections over a three-hour period [20].
6. On two rescue flights from Europe to Korea, several of the passengers had primary Covid-19 infections. However, the passengers wore N95-masks, which are considerably more effective in preventing transmission than the masks used now on US domestic flights. (In the *Lancet* paper, the point estimate of effectiveness for N95 masks is 96%.) Nonetheless, a single secondary infection did arise on each flight, one of which is probably tied to removal of the mask during a visit to the toilet. See Bae et. al. [21]
7. On a flight from Guangzhou to Toronto, two passengers traveling together had primary Covid-19 infections [22]. They were wearing masks, as were other passengers. There were no known secondary infections on the flight.

Of course, the flights described above are not a random sample of flights performed. If no contagious people are on board, then there are obviously no secondary infections. But *τ*_0_ is only relevant when contagious passengers do board and when, furthermore, masks fail to prevent them from transmitting the virus. The probability that this adverse combination of circumstances arises is *QQ*_*M*._

These flights offer varying levels of information and divergent estimates about *τ*_0_. The paper by Hertzberg and Weiss about SARS found that most on-board infections arose more than two rows from the contagious person. We did not work with that article because its authors were lead authors of the later PNAS paper, which effectively superseded the original conclusion about spread. (In the latter paper, they noted that mingling in the boarding area for the SARS flight might explain why some passengers with secondary infections were seated far from those with primary infections.) The absence or near-absence of infections in the flights discussed in (6) and (7) is not illuminating about *τ*_0_ : Even if that parameter was high, mask usage would have been expected to preclude transmission. The flight discussed in [20] generated Hertzberg and Weiss ‘s estimate of a transmission probability of .0045 per minute. However, Moser et al report that the airplane’s ventilation system was not working during the exposure period, meaning that .0045 would be an upper bound for *τ*_0_—and probably a high upper bound--on planes with functioning air filters. (The HEPA air filters on jets like the Boeing 737 and the Airbus 320 are highly effective, as US airlines point out. However, the examples above show that while these filters reduce viral transmission, they cannot completely prevent it.)

For the other flights listed above, we estimated *τ*_0_ approximately to achieve a match with the observed number of secondary infections, given the flight duration and generally assuming our mid-range estimates for *ω and λ*. For the Sydney-Perth flight, the secondary infection rate of 40% (8 out of 20) suggests a *τ*_0_-value of about .002. The 92% rate of nearby secondary infections on Vietnam Flight 54 implies that *τ*_0_ was .0.05 if not higher (and, even then, the damping of risk with distance (*ω*) would have to be closer to the lower bound in the *Lancet* paper (i.e. a drop of about 8% per meter) than to their point estimate (about 50% per meter)). On the Tel Aviv-Frankfurt flight, by contrast, the 17% secondary infection rate for nearby passengers would yield *τ*_0_−estimate somewhere between .0005 and .002, depending on how one cumulates the risk posed by several passengers with primary infections seated next to one another.

How does it all add up? Reflecting this range of *τ*_0_−estimates and a dose of “engineering judgment,” we advance another triangular distribution, in the range from 0 to .006:

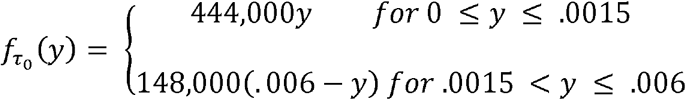

This distribution has a median of .0015 and a mean of .0019.

In summary, our distributions for key parameters are:

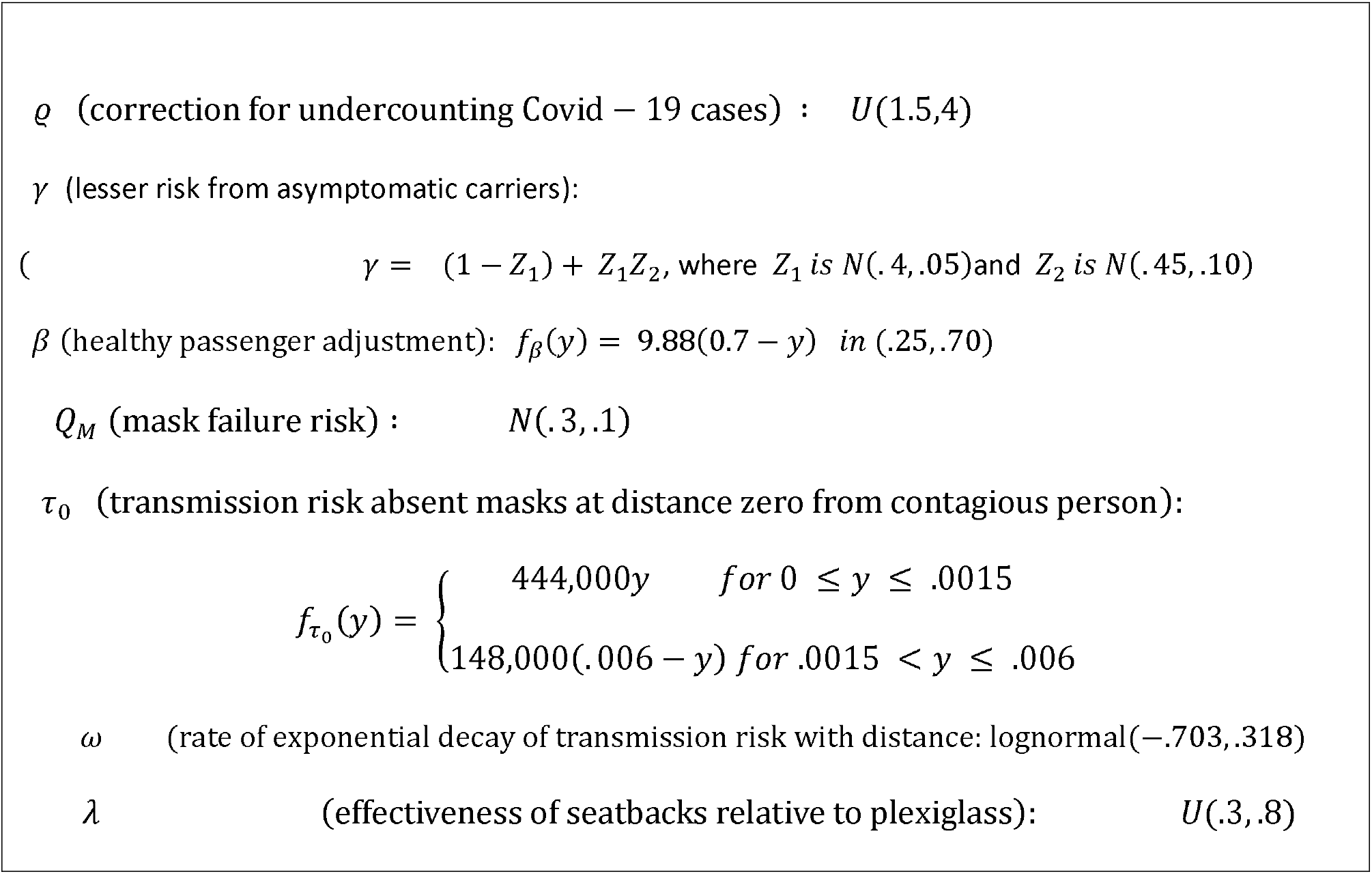

### IV. Results

Given the number of random variables in the expression for P and their differing distributions, the distribution for P does not take any familiar analytic form. It is necessary to use simulation to investigate the behavior of P, and we did so, treating the various random processes as independent. In nearly all respects, an independence assumption is highly plausible: the degree to which confirmed Covid-19 cases underestimate the true number bears little relation to the effectiveness of seatbacks relative to plexiglass. We did note that, for Vietnam Flight 54, a high estimate of *τ*_0_ would accompany a value *ω* of that yields slow damping of risk with distance. But this example of possible correlation is an exception even to the overall information about *τ*_0_ and *ω*. An independence assumption is viable even if not perfect.

We conducted 10,000 simulations, assuming an all-coach configuration of the aircraft with 29 rows and 174 seats. We assumed that a Covid-19 contagious person is equally likely to be in any of those seats when the flight is full and, when the plane is full except for middle seats, would be equally likely to take any of the 116 (i.e., 174*(2/3)) available seats. The pattern of infection depends slightly on whether the contagious person is in a window, middle, or aisle seat, and contagious people in the first two rows or back two rows of the plane have fewer vulnerable passengers within two rows of them. All these considerations entered into the simulations and calculations.

Figure 1 offers histograms of the simulation results, while Table 2 presents various percentiles of the distributions for the probability of contracting Covid-19 on a US domestic flight two hours long. It also shows the mean and standard deviation of the 10,000 risk outcomes. The unbiased point estimates are the means of the distributions for “all seats full” and “middle seats empty,” which were 1 in 3928 and 1 in 6423, respectively, differing by a factor of 1.64. Because the two risk estimates depend on the various parameters in essentially the same way, it is unsurprising that the ratio of risk between the two policies was largely the same at each percentile, with slight variation around 1.64.

**Table 2:** Results of 10,000 Simulations about Risk of Contracting Covid-19 on a Two-Hour US Domestic Jet Flight

| Distribution of Estimated Infection Probability |  |  |
| --- | --- | --- |
| <u>Percentile</u> | <u>Fill All Seats</u> | <u>Middle Seats Empty</u> |
| 2.5<sup>th</sup> | 1 in 33,681 | 1 in 56,754 |
| 10<sup>th</sup> | 1 in 14,590 | 1 in 24,319 |
| 25<sup>th</sup> | 1 in 8306 | 1 in 13,824 |
| 50<sup>th</sup> (Median) | 1 in 4845 | 1 in 7,994 |
| 75<sup>th</sup> | 1 in 2980 | 1 in 4878 |
| 90<sup>th</sup> | 1 in 1992 | 1 in 2980 |
| 97.5<sup>th</sup> | 1 in 1313 | 1 in 2112 |
| Mean of Estimate: | 1 in 3928 | 1 in 6423 |
| Standard Deviation of Estimate: | 1 in 5215 | 1 in 8370 |
| Coefficient of Variation | 0.753 | 0.767 |

**Figure 1:**
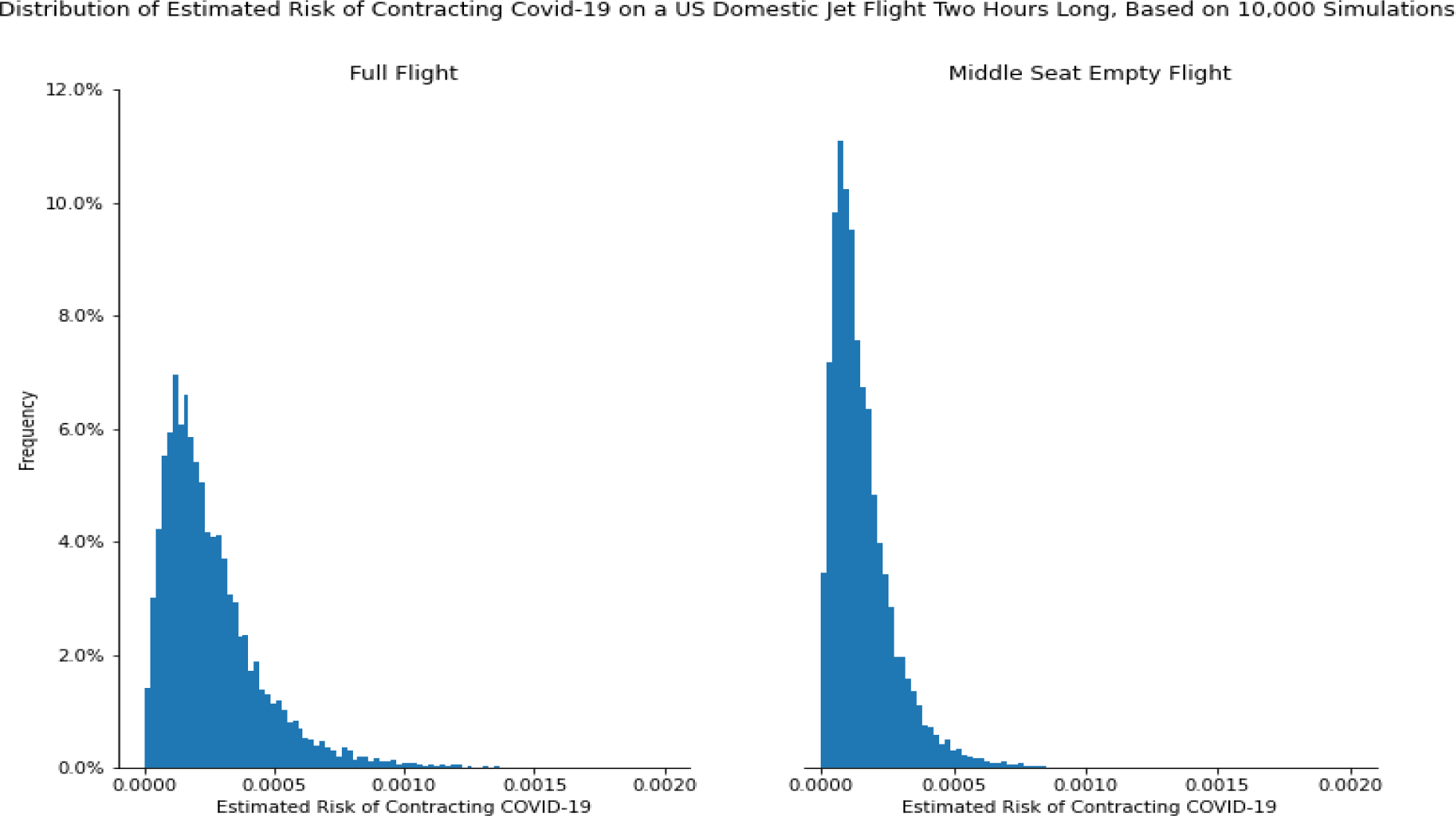
Distribution of Estimated Risk of Contracting Covid-19 on a US Domestic Flight Two Hours Long, Based on 10,000 Simulations

A risk multiplier of 1.64 for a flight with all seats full versus one with empty middle seats makes intuitive sense. Having five other passengers in one’s row rather than three would seem to raise by a factor of roughly 5/3 = 1.67 the chance that one of them is contagious. In adjacent rows, having six passengers rather than four raises the corresponding likelihood by a factor of 1.5. Thus, one would anticipate that, even when the specifics of distance are considered, the risk multiplier would not stray too far from 1.6. And it does not.

Table 3 notes that transmission risk is lowest when the contagious passenger is in a window seat (A or F), and highest when she is in an aisle seat (C or D). This pattern makes sense: someone in seat C is closer than someone in A to the passengers in seats D, E, and F of the same row, and both these passengers are equidistant from a traveler in seat B. Moreover, the distance from A to C is the same as that from C to A. Hence, the grid distances to the five uninfected passengers in the same row are lower from seat C than from seat A. The same considerations apply in connection with the row ahead and the row behind.

**Table 3:** Infection Probability and the Location of a Contagious Passenger

| Contagious Person's Seat | Average Infection Probability for Other Passengers** |  |
| --- | --- | --- |
|  | <u>Fill All Seats</u> | <u>Middle Seat Empty</u> |
| Window (A or F) | 1 in 565 | 1 in 920 |
| Middle (B or E) | 1 in 485 | ---- |
| Aisle (C or F) | 1 in 451 | 1 in 726 |

\*\* Contingent on actually having such a contagious passenger, and on universal mask usage. On most flights, there are no contagious passengers.

To end the risk calculation, we divide through by the number of uninfected passengers who boarded the flight. Suppose for example that one contagious traveler comes on board, and generates an expected 0.35 new infections among the 173 other passengers despite mask usage.. Then the chance that a random-chosen passenger among those 173 comes down with Covid-19 would be 0.35/173. The risk would be higher for those close to the contagious passenger and virtually zero for those many rows away, but the risk for the randomly-chosen passenger is given by (1/173) times the sum of risks across all seats (which is the expected number of secondary infections).

Of course, the distribution of Covid-19 risk has substantial spread. For both “fill all seats” and “middle seats empty”, the interquartile range from the 25^th^ percentile to the 75^th^ percentile is 95% as large as the median, and the standard deviations of outcomes are about 75% of the means. The 97.5^th^ percentiles of risk are about a factor of three above the point estimates, while the 2.5^th^ percentiles are about a factor of eight below. Such a sizable level of uncertainty was unavoidable, given the lack of precise information on matters like as the effectiveness of seatbacks against transmission, the extent to which confirmed Covid-19 cases understate all cases, and the degree to which air travelers are less likely to carry contagious Covid-19 than the citizenry as a whole. Still, the analysis seems decisively to rule out possibilities like a one-in-a-million chance of catching Covid-19, or a one-in-ten risk of doing so.

## Discussion

Aviation officials often assert that many everyday activities pose greater Covid-19 risks than those on airplanes, and that statement is true. But it is doubtful that two hours on a domestic flight is as safe as two typical hours on the ground. In the US in week from 9/17/20 to 9/23/20, an average of 36,000 Covid-19 infections were confirmed per day. Using a scaling factor of 2.75 (i.e., our mean estimate for *ρ*), the estimated true number of new infections each day was 99,000. In a US population of 330 million, the daily infection probability would be 99,000/330 million, which is 1 in 3333. Assuming 16 waking hours and no new infections during sleep, the chance of infection over a two-hour waking period would be approximately (2/16)*(1/3333) = 1 in 26,700. And given that our “healthy traveler” parameter *β* has a mean of 0.4, the corresponding risk over two hours would 1 in 66,800 for air passengers. That risk level falls below the 2.5^th^ percentile for infection risk even when middle seats are kept empty.

The main reason that Covid-19 is so fearsome is that it entails a risk of death. Thus, we should convert estimates of in-flight infection risk to estimates about subsequent deaths that these infections cause. The passengers at greatest risk are those above age 65 and those with major medical conditions, groups that would seem less likely to fly out of fear of Covid-19. Thus, we might assume that the mortality risk for domestic air passengers who contract Covid-19 is 0.1%, well below the US population-wide average of 0.5% to 0.7%. However, covid-19 infections on planes can cause deaths to some people who were not passengers (e.g., a 22-year traveler gets infected, and passes the virus on to his elderly grandparents). These indirect victims of infections incurred during flights (i.e. of tertiary infections) could well outnumber the direct victims.

A quantity familiar in this pandemic is R_0,_ the average number of new infections generated directly by an infected person. The quantity E(Further Inf), the mean total number of further infections that person causes, follows:

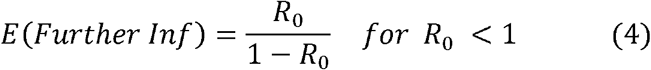

Using the conservative estimate that *R*_0_ = 0.5, the quantity *E*(*Further Inf*) = 1 under (4). Assuming a 0.6% death risk for people indirectly infected because of in-flight transmission of Covid-19, the expected total number of deaths per in-flight infection would be 0.001 + 1*.006 = .007. Thus, if a given passenger on a full flight has a 1 in 3,928 chance of getting infected, the resulting number of deaths would on average be about (1/3928)*.007 = 1 per 561,000 passengers. Under these assumptions, the vast majority of those deaths would befall people who were not even on the plane.

On flights that are 66.7% full under middle seat empty, the death toll would fall to 1 per 918,000 passengers. Moreover, even an airline willing to fill every seat does not expect to do so: it presumably aspires to a passenger load factor (passenger miles divided by seat miles) around 85.1%, which prevailed in 2019 on US flights. Having 85.1% of seats taken is consistent with roughly 55% of middle seats full and 45% empty. With that load factor, secondary Covid-19 infections would yield an effective death risk under “fill all seats” of about 1 per 710,000 passengers rather than 1 in 561,000.. Airlines that keep middle seats empty could well try to fill nearly all of them, so their mortality risk would remain about 1 in 918,000. All these estimates are considerably higher than the risk of perishing in a US air crash unrelated to Covid-19, which is about 1 in 34 million [10]).

### VI. Final Remarks

The key determinants of in-flight infection risk are the mean number of contagious people on board, the cloth/surgical mask success rate, and the expected number of infections generated per contagious person. For a given mean number of infections, the distribution of risk across uninfected travelers is irrelevant: another distribution with the same mean that reduced risk in 15B but increased it in 15D would yield the same estimate for *P*. Similarly, the general approach is valid whether one estimates the risk to a given passenger through accumulating risks per minute or via another approach tied to total dose absorbed over the flight. Thus, the procedure advanced here to estimate P can accommodate various modifications to its assumptions.

Because confirmed Covid-19 infections on US jet flights have been very rare, the estimates offered here about in-flight infection risk might provoke skepticism. But actual load factors on such flights have been so low in recent months that the risk estimates calculated here would not apply. In months to come, however, load factors under drastically-reduced air schedules could well approximate those we have considered. Thus, our estimates might overestimate what happened in the past, but they could be highly relevant to the future.

More fundamentally, there have been over 8 million confirmed Covid-19 infections in the US, and perhaps 14 million more that were never confirmed. Despite this circumstance, US follow-up processes have generally been so weak that we have scant information about how many infections arose in what settings. How many Americans have become infected while riding on buses? In bowling alleys? The only cases we know much about are “superspreader” events in which large numbers of people simultaneously come down with Covid-19. But airplane transmission by its localized nature would not produce superspreader events. We do not usually know which passengers with primary infections boarded airplanes. We know even less about which passengers sustained secondary infections, and we know practically nothing about tertiary infections that ultimately trace back to aircraft cabins. (Contact tracing has been far superior in some Asian countries, but they have so few Covid-19 cases that their Q-values on flights are orders of magnitude below those in the US.)

The calculations here, while hardly exact, do suggest a measurable reduction in Covid-19 risk when middle seats on aircraft are deliberately kept open. Relinquishing 1/3 of seating capacity to achieve that reduction is a high price to pay. But so great is the reluctance to fly amid the US pandemic that steps that reassure the public (e.g., flying extra sections of flights when demand exceeds 2/3 of capacity) might well increase revenue more than they increase costs.

## Data Availability

All data are readily available in public sources, like new confirmed Covid-19 cases by day in US states. These sources are identified in the paper.

## Acknowledgements

To be supplied

## Notes

### Competing Interest Statement

The authors have declared no competing interest.

### Author Declarations

No such body has considered the work.

### Summary of Updates

The revision is more explicit about the assumptions underlying the calculations, and is tied more closely to the rapidly evolving literature about the risks that Covid-19 poses to air travelers.

## References

[1] I. Duncan, “Nearly 11,000 People Have Been Exposed to the Coronavirus on Flights, the CDC Says,” Washington Post, September 19, 2020, available at https://www.washingtonpost.com/local/trafficandcommuting/nearly-11000-people-have-been-exposed-to-the-coronavirus-on-flights-the-cdc-says/2020/09/19/d609adbc-ed27-11ea-99a1-71343d03bc29_story.html

[2] D. Freedman and A. Wilder-Smith, “In-flight Transmission of SARS-CoV-2: A Review of the Attack Rates and Available Data on the Efficacy of Face Masks,” Journal of Travel Medicine, September 25, 2020 available at https://academic.oup.com/jtm/advance-article/doi/10.1093/jtm/taaa178/5910636

[3] A. Byrne et. al.(2020), “Inferred Durations of Infectious Period of SARS-Cov-2: Rapid Scoping Review and Analysis of Available Evidence for Asymptomatic and Symptomatic Covid-19 Cases,” available at https://www.medrxiv.org/content/10.1101/2020.04.25.20079889v1.full.pdf

[4] Y. Ling et. al.(2020), “Persistence and Clearance of Viral RNA in 2019 Novel Coronavirus Disease Rehabilitation Patients,” Chinese Medical Journal 133(9), pp. 1039–1043, May 5, 2020, available at https://www.ncbi.nlm.nih.gov/pmc/articles/PMC7147278/

[5] F. Havers et al, “Serioprevalence of Antibodies to Sars-Cov-2 in 10 Sites in the United States, March 23-May 12, 2020, JAMA Internal Medicine, July 21, 2020, available at https://jamanetwork.com/journals/jamainternalmedicine/fullarticle/2768834

[6] D. Chu et. al.(2020) “Physical Distancing, Face Masks, and Eye Protection, To Prevent Person-to-Person Transmission of SARS-Cov-2 and COVID-19: A Systematic Review and Meta-Analysis, The Lancet, 395(10242), pp. 1973–1987, June 27, 2020

[7] V.S. Hertzberg, H. Weiss, et. al. (2018) “Behaviors, Movements, and Transmission of Droplet-Mediated Respiratory Diseases During Transcontinental Airline Flights, Proceedings of the National Academy of Sciences,115(14), pp. 3623–3627, April 3, 2018.

[8] Worldometer.com “Age, Sex, Existing Conditions of COVID-19 Cases and Deaths,” data for confirmed cases by day for the United States, available at https://www.worldometers.info/coronavirus/coronavirus-age-sex-demographics/

[9] G. Meyerowitz-Katz and L. Merone “A Systematic Review and Meta-analysis of Published Research Data on Covid-19 Infection-Fatality Rates,” July 7, 2020, available at https://www.medrxiv.org/content/10.1101/2020.05.03.20089854v4

[10] Recovery Trial Network, “Low-cost Dexamethasone Reduces Death by Up to One-Third in Hospitalized Patients with Severe Respiratory Complications of Covid-19” June 16, 2020, available at https://www.recoverytrial.net/news/low-cost-dexamethasone-reduces-death-by-up-to-one-third-in-hospitalised-patients-with-severe-respiratory-complications-of-covid-19

[11] R. A. Armstrong, A. D. Kane, and T. M. Cook, “Outcomes from Intensive Care in Patients with Covid-19: A Systematic Review and Meta-analysis of Observational Studies,” Anaesthesia, June 30, 2020, available at https://associationofanaesthetists-publications.onlinelibrary.wiley.com/doi/full/10.1111/anae.15201

[12] D. Oran and E. Topol, “Prevalence of Asymptomatic SAR-CoV-2 Infection; A Narrative Review,” Annals of Internal Medicine, September 2020, available at https://www.acpjournals.org/doi/10.7326/M20-3012

[13] CSPEC (2020), “Defining High-Value Information for Covid-19 Decision Making,” available at https://www.medrxiv.org/content/10.1101/2020.04.06.20052506v1.full.pdf

[14] Y. Wang, et.al. “Reduction of secondary transmission of SARS-CoV-2 in households by face mask use, disinfection and social distancing: a cohort study in Beijing, China,” BMJ Global Health, 5(5),2020, available at https://gh.bmj.com/content/5/5/e002794

[15] S. Eikenberry, M. Mancuso, E. Iboi, T. Phan, K. Eikenberry, Y. Kuang, E. Kostelich, and A. Gumel, “To mask or not to mask: modeling the potential for face mask use by the general public to curtail the COVID-19 pandemic,” Infectious Disease Modeling, vol. 5, pp. 293–308, 2020.

[16] V. Hertzberg and H. Weiss, “On the 2-Row Rule for Infectious Disease Transmission on Aircraft,” Annals of Global Health, 82(5), September-October 2016, pp. 819–823.

[17] N.C. Khanh et. al. “Transmission of Severe Acute Respiratory Syndrome Coronavirus 2 During Long Flight, Emerging Infectious Diseases, 26(11), November 2020, available at https://wwwnc.cdc.gov/eid/article/26/11/20-3299_article#tnF2

[18] S. Hoehl et. al. “Assessment of SARS-CoV-2 Transmission on an International Flight and Among a Tourist Group,” JAMA Network Open, available at JAMA Network Open. 2020;3(8):e2018044. doi:10.1001/jamanetworkopen.2020.18044

[19] H. Speakes et. al., “Flight-Associated Transmission of Severe Acute Respiratory Syndrome Coronavirus 2 Corroborated by Whole Genome Sequencing, Emerging Infectious Diseases,” 26(12), December 2020, available at https://doi.org/10.3201/eid2612.203910

[20] M.R. Moser et. al., “An Outbreak of Influenza aboard a Commercial Airliner,” American Journal of Epidemiology, 110(1), pp. 1–6, 1979

[21] S. H. Bae et. al., “Asymptomatic Transmission of SARS-Cov-2 on Evacuation Flight, Emerging Infectious Diseases, 26(11), November 2020, available at https://wwwnc.cdc.gov/eid/article/26/11/20-3353-f1\

[22] K. Schwartz et. al. “Lack of Covid-19 Transmission on an International Flight,” CMAC 192(5), April 14, 2020, available at https://www.ncbi.nlm.nih.gov/pmc/articles/PMC7162437/

[23] A. Barnett (2020), “Aviation Safety: A Whole New World?” Transportation Science 54(1), pp. 84–96, January-February 2020

